# The role of vaccination and public awareness in medium-term forecasts of monkeypox incidence in the United Kingdom

**DOI:** 10.1101/2022.08.15.22278788

**Authors:** SPC Brand, M Cavallaro, J Hilton, LM Guzman-Rincon, T House, MJ Keeling, DJ Nokes

## Abstract

Monkeypox virus (MPXV) is spreading rapidly through close human-to-human contact primarily amongst communities of men-who-have-sex-with-men (MSM). Behavioural change arising from increased knowledge and health warnings may decelerate the rate of transmission and Vaccinia-based vaccination is likely to be an effective longer-term intervention. Here we investigate the current epidemic within the UK population and simulate control options over a 12 week projection using a stochastic discrete-population transmission model which includes MSM status, rate of formation of new sexual partners, and an underlying random sized metapopulation structure. We find that the virus may have already infected a significant proportion of the MSM group with the highest sexual activity (32.5%; 15.9% - 44.9% prediction IQR); the associated immunity, albeit among groups that form a small but sexually active part of the MSM community in the UK, coupled to behavioural driven decrease in the transmission rate of individuals infected with monkeypox, leads to case incidence flattening and then declining over the projection period (12 weeks). Vaccination is most beneficial when targeted to MSM with highest activity if delivered in the near term to further interrupt transmission amongst those driving the epidemic.

## Introduction

There has been sustained incidence of monkeypox virus (MPXV) in European and North American countries since May 2022, leading to a WHO declaration of a public health emergency of international concern on 23rd July 2022 [1]. It is likely that vaccines developed to target smallpox have reasonable efficacy against MPXV-induced disease [2–4], and given the significant skew in cases towards men-who-have-sex-with-men (MSM) it has been suggested that vaccines developed against MPXV should be offered as pre-exposure prophylactic (PrEP) targetted at this group [5]. Towards this goal the UK Health Security Agency (UKHSA) and the Joint Committee on Vaccination and Immunisation (JCVI) have recommended the use of the Modified Vaccinia Ankara (MVA) smallpox vaccine Imvanex (called Jynneos in the USA) for at-risk groups for monkeypox in the UK [6].

monkeypox predominantly spreads from person-to-person through physical contact with the infectious rash, scabs and/or fluids of infected individuals [7]. Traditionally, MPXV incidence has been sporadically observed in sub-saharan Africa following zoonotic spread from wildlife reservoirs and with uncommon person-to-person transmission being associated with household cohabitation [8]. However, as background immunity to MPXV due to mass smallpox immunisation has waned there has been a substantial increase in monkeypox incidence in some sub-Saharan African countries [9,10].

In the recent outbreak in Europe and North America, the epidemiology of MPXV has skewed towards a higher case frequency among MSM people compared to non-MSM groups. This is in contrast to the traditional epidemiology of MPXV described above, and it is possible that MPXV has found a niche in high income countries (HICs) among individuals with high frequencies of physical contact. It is also suggestive that public awareness of monkeypox symptoms, and thereby reducing onward transmission during the symptomatic phase of an infectious episode, could be effective at limiting the overall impact of MPXV. Early epidemiological modelling of the transmission potential of MPXV in HICs has identified the potential for MPXV to spread among sexual contact networks due to the assortative mixing of individuals with highly frequent sexual encounters, with the distribution of the number of encounters best described by a heavy-tailed distribution [11]. This early concern seems to have been confirmed, whereas early detailed individual-based transmission modelling for MPXV without a focus on sexual contact network dynamics has been proven over-optimistic [12].

In this simulation study, we use a bespoke MPXV transmission model calibrated to both the social structure and demography of the United Kingdom, and reported weekly MPXV incidence, to make projections of future MPXV incidence over a medium-term time horizon (12 weeks ahead). We use robust Bayesian inference to make model parameter inference (see **Methods** and **Supporting information**) as well as validating the model’s sequential forecast accuracy (see **Supporting information**). The MPXV transmission model is capable of exploring the potential impacts of vaccinating the most at-risk MSM people against MPXV and reduction in transmission due to self-imposed reductions in physical contacts whilst having symptoms. The purpose of this study is three-fold: First, to forecast the likely scale of total MPXV incidence over the next few months in the United Kingdom both in terms of posterior predictive expectation and posterior inter-quartile range (IQR). Second, to assess the evidence for a reduction in transmission per case having already occurred. Third, to consider the scale of the MPXV incidence conditional on three pessimistic scenarios occurring. The pessimistic scenarios considered, ordered by our *a priori* belief on their likelihood occurrence are:

- **No further behavioural response**. Any reduction in transmission due to awareness of MPXV symptoms ceases when projecting forwards.
- **No/ineffective vaccination rollout**. The vaccination campaign in the UK is ineffective due to some unforeseen factor, e.g. vaccine hesitancy in most at-risk groups.
- **Reasonable worst case scenario**. Both pessimistic scenarios above occur.

## Results

We infer that the on-going high ratio of MSM cases compared to non-MSM cases observed in the UK is due to MPXV having a low transmission potential outside high frequency sexual contact groups; the reproductive ratio for other transmission pathways on 1st May 2022 was estimated to be 0.47 (0.054, 1.1; 95%CI) (Table S1 for all parameters and Fig. S2 for visualisation of posterior distributions), whereas the risk of transmission per sexual contact on 1st May 2022 was inferred to be 31% (15%, 52%; 95%CI) - suggesting the reproductive ratio will be greater than one within populations that have more than 3 sexual contacts during the 2-week infectious period (Table S1). We find evidence that the risk associated with both transmission pathways decreased over June and July 2022. By the WHO announcement of a public health emergency of international concern (23rd July 2022) we estimate that the reproductive ratio for other transmission pathways had decreased to 0.07 (0.003, 0.20; 95%CI; posterior probability of >10% decrease compared to 1st May was 0.995) and the effective risk of transmission per sexual contact had decreased to 20.7% (9.2%, 35.7%; 95%CI; posterior probability of >10% decrease compared to 1st May was 0.9) (Fig. 1).

**Figure 1.**
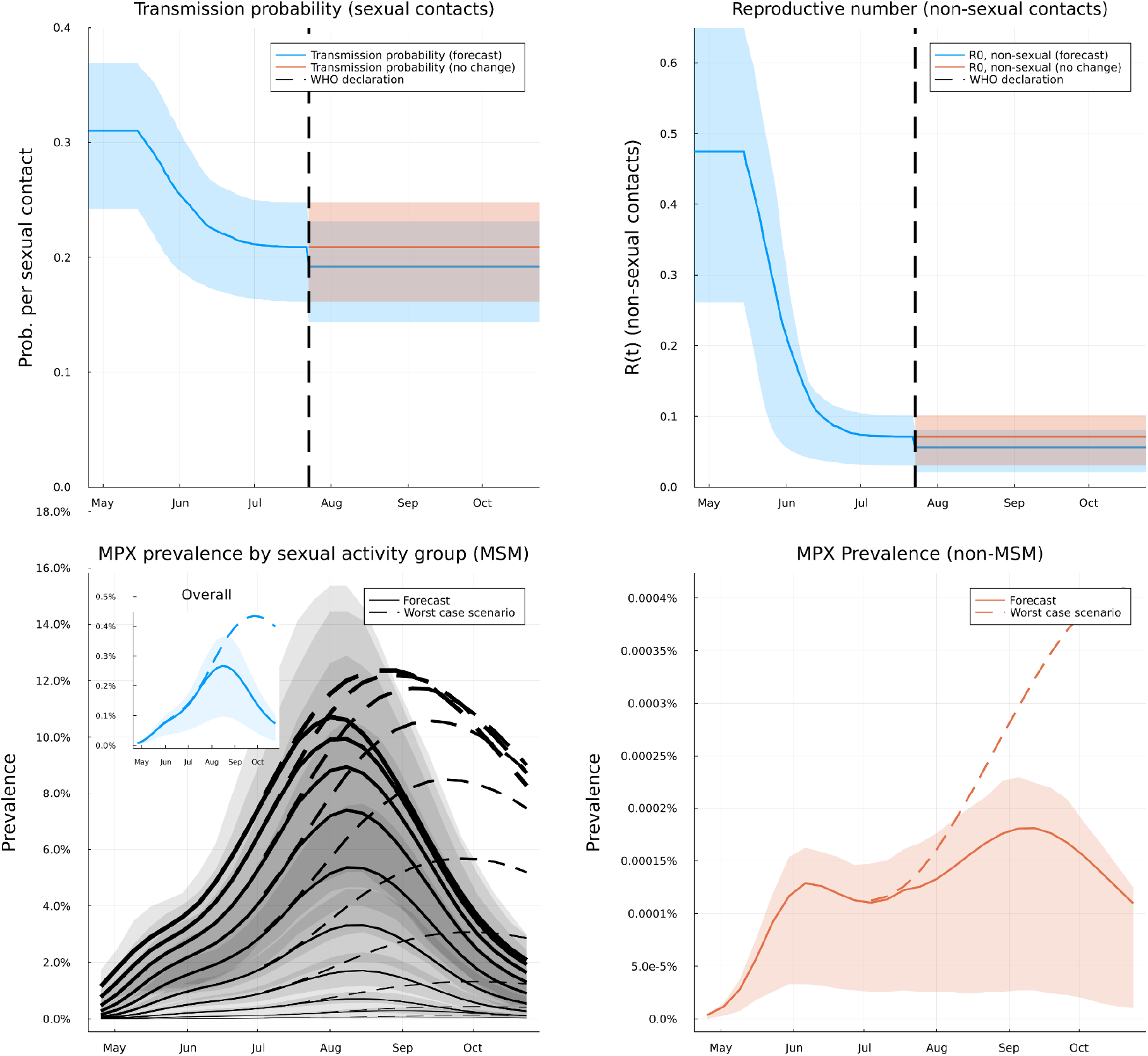
Temporal trends in MPX effective transmissibility and prevalence. Posterior predictions for the probability of transmission per sexual contact over time (*top left*) and background reproductive number (*top right*). Posterior mean value for forecast scenario (risk per contact decreases after WHO announcement of public health emergency of international concern; blue curve) and no change further scenario (red curve). Posterior predictions for monkeypox prevalence among MSM sexual activity groups (*bottom left*), overall sexually active MSM people (*bottom left inset*), and the overall non-MSM and sexually inactive MSM population (*bottom right*). Among MSM sexual activity group prevalence among least (thinnest black curves) to most (thickest black curves) sexually active groups (see Methods for group definitions). All background shading shows posterior IQRs (25-75% posterior prediction intervals).

We interpret the decreased transmission potential of MPX as being due to greater public awareness of monkeypox disease and, therefore, physical contacts that would have occurred with individuals with MPXV symptoms prior to greater awareness have been more frequently avoided in May-July 2022. We consider it *a priori* likely that the WHO announcement will further decrease the effective transmissibility of MPXV, based on public behavioural response to WHO announcements in the early stages of the COVID-19 pandemic [13], although our prior belief based on personal communication with UKHSA is that this effect is small compared to any reduction in transmission potential prior to the WHO announcement. The level of transmission within the high-risk MSM groups is likely to be further limited by immunisation. In the UK, the first MSM believed to be at high risk of MPX exposure volunteered for a dose of the Imvanex/Jynneos vaccine on 16th/17th July, with current projections suggesting ∼5000 doses offered in London per week throughout August [14].

Our default MPX medium-term (12 week lookahead) projection assumes that the WHO announcement triggered a further decrease in MPX effective transmissibility which we infer, albeit on limited data (see Fig. 1) and that the projected rate of vaccine uptake will be realised and sustained (Fig. 5). In the following we give projections based on the posterior mean over simulations after parameter inference, as well as reporting the posterior median and posterior inter-quartile range (IQR). This forecast implies that, in the week starting 15th Aug 2022, actively infectious MPX prevalence in the UK will be 0.30% among MSM (0.094%, 0.22%, 0.42% IQR) and 0.00017% among non-MSM (0.000081% median 0.000024%-0.00021% IQR; Fig. 1). This headline projection of peak prevalence among MSM individuals hides substantial variation within the MSM community, with the most sexually active group of MSM people predicted to have peaked in the week starting 1st Aug 2022 with prevalence of 11.4% (10.3% median, 5.0%-16.4% IQR; Fig. 1). Under our reasonable worst case scenario (no further decrease in effective transmissibility and an ineffective vaccination campaign) we forecast that MPX prevalence will continue to grow among MSM and non-MSM until the third week in September and peak at substantially higher prevalence (Fig. 1). In the worst case scenario, the limiting factor for MPX spread is depletion of susceptible highly sexually active individuals due to unconstrained spread (Fig. 1) - this means that the relative difference between the default and reasonable worst case assumptions is greatest in the less sexually active groups and the non-MSM population.

Our medium-term (12 week lookahead) projection is that weekly MPX case incidence will peak in the week starting 22nd August with 451 (327 median, 143 - 612 IQR) cases among MSM and 21 (10 median, 3 - 26 IQR) cases among non-MSM. We then forecast that weekly case incidence will trend downwards in September 2022 with reported cases among MSM forecast: 416 (304 median, 140 - 579 IQR) on first week starting in September, and 218 (156 median, 69 - 299 IQR) on first week starting in October. Forecast numbers for non-MSM are 25 (12 median, 3 - 31 IQR) on the first week starting in September, and 23 (10 median, 3 - 28 IQR) on the first week starting in October (Fig. 2). We project cumulative MPX cases up until the week starting 24th October being 6,341 (5,574 median, 4,048 - 7,758 IQR) and 349 (226 median, 136 - 423 IQR) among non-MSM (Fig. 2).

**Figure 2.**
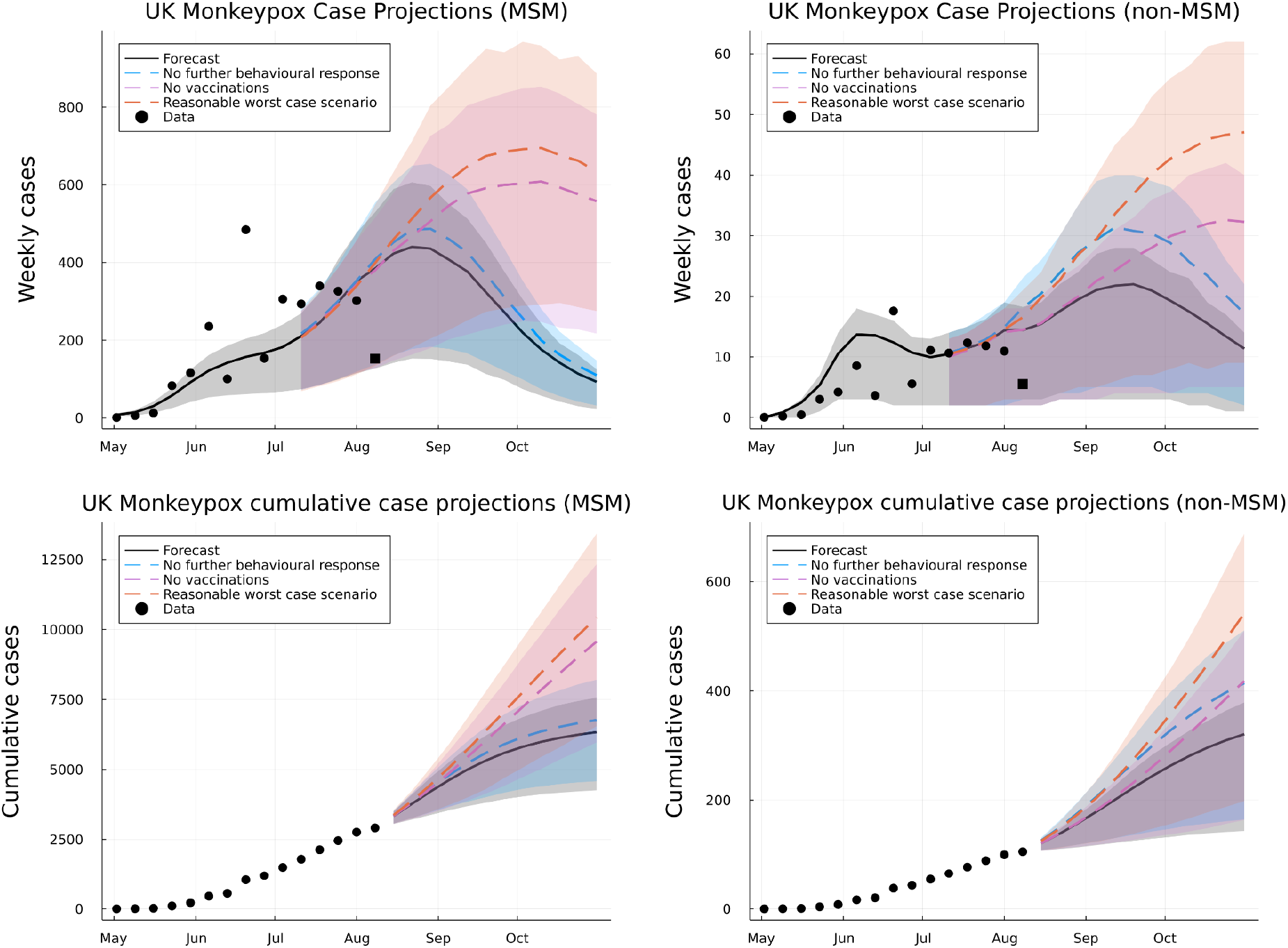
Forecasts and scenario projections for weekly monkeypox incidence. Posterior predictions over a 12 week horizon for weekly case incidence among MSM (*top left*) and non-MSM (*top right*) and cumulative case incidence among MSM (*bottom left*) and non-MSM (*bottom right*). Four scenarios are shown: forecast scenario (black solid curve), “no further behavioural response” scenario (blue dashed curve), “No/ineffective vaccination” scenario (purple dashed curve), “reasonable worst case” scenario (red dashed curve). Data used in model inference is shown against predictions (black circles) as well as most recent week hold-out data (black squares). All curves represent posterior mean predictions, background shading showing posterior IQRs (25%-75% prediction intervals).

However, as mentioned above, this forecast is contingent on our *a priori* belief that effective transmission of MPX will continue to fall. The more pessimistic plausible scenarios, outlined above, predict worse MPX incidence than the forecast projection. In the **No further behavioural response** scenario we project that MPX cases will peak at a marginally higher rate with marginally more cases over the 12 week projection horizon compared to the default scenario: MSM cases 6,814 (6,070 median, 4,403 - 8,247 IQR), non-MSM cases 458 (295 median, 164 - 558 IQR; Fig. 2). The similarity between the forecast scenario and the **No further behavioural response** scenario is due to our prior belief that the WHO announcement has not generated a substantive further change in behaviour. In the **No/ineffective vaccination rollout** scenario we project that the weekly incidence continues trending upwards until early October and that there are substantially more MPX cases over the 12 week projection horizon compared to the forecast scenario: MSM cases 9,193 (8,296 median 5,441 - 11,772 IQR), non-MSM cases 436 (275 median, 151 - 546 IQR; Fig. 2). This projection implies that rapidly getting to 7,500 imvanex first doses per week taken up by individuals with typically more than one new sexual partner a month, our forecast projection, reduces the posterior median projection of MPX cases by 17.7% over a 12 week horizon compared to no/ineffective vaccination rollout. In the **Reasonable worst case** scenario we projected similar, but higher, weekly incidence trends to the no vaccination scenario: over a 12 week horizon total MSM cases 10,112 (9,152 median, 6,145 - 12,883 IQR), total non-MSM cases 586 (377 median, 192 - 776 IQR).

## Discussion

We developed a novel stochastic discrete population model to enhance understanding and make projections on MPX incidence in the UK. The epidemiology of MPX in the UK suggests a network-based modelling approach, but this is both challenging to develop and to make inference against the currently available data. Our relatively simple model aimed to capture much of the essential features of a transmission network, in particular differential behaviour, whilst being feasible to fit and re-fit to available data streams, hence providing a rapid projection tool providing results of immediate benefit to policy makers. Our work is partly inspired by useful approximations of individual-scale models by metapopulation models [15], and we note that as of mid-July greater than 70% of the determined MPX incidence in the United Kingdom has been in London [16]. Whilst we don’t explicitly fix the metapopulation size structure used in this study (see Methods); the model we have developed is flexible and capable of recreating typical features of metapopulation dynamics, such as a concentration of cases in one group.

Our investigation leads us to believe that from the first couple of weeks in August MPX incidence is flattening and soon declining with current cumulative case numbers (∼2600) increasing to ∼6300 (posterior mean) / ∼5500 (posterior median) over the next 12 weeks (up until week starting 24th October 2022). We predict an already high prevalence among the small number of people in the most sexually active groups, which has an effect of limiting the size of the epidemic going forwards. The model-based inference on the epidemic trajectory suggests that transmission potential per infected person (either MSM/non-MSM) has already decreased significantly (by approximately 30%) since start of the outbreak, which we interpret as the effect of public awareness of the existence/threat of MPX and its symptomatic identifiability. If projected vaccination rates of 7-8000 doses per week are achieved among the MSM with highest sexual frequency of new partners (see Methods for sexual activity group definition) we forecast a substantial number of cases will be averted over next 12 weeks (∼17.8% compared to no/ineffective vaccination). However, there are significant risks at the margin of worse outcomes in the medium term, such as seeing a total of > 10k cases by the end of October, if a pessimistic scenario occurs, e.g. a failure of the vaccination programme against monkeypox.

The simplicity of the model and weaknesses of the epidemiological data lead to limitations of our work. The delay between symptom onset, seeking treatment, case confirmation and reporting has changed over the course of outbreak, for example, onsets across April were included in the first week of May reporting, which could influence inference. Additionally, it is not possible using this model to infer a changing trend in probability of detecting infected individuals, for example if the case ascertainment for monkeypox has been falling since June this model could erroneously attribute the slowing case rate to behavioural change. In mitigation we use a robust Bayesian inference method and the longer delayed reporting occurred early in the outbreak when at small numbers, essentially, trading decreased likelihood of radically incorrect inference based on overfitting to outliers for wider prediction intervals. We also assessed the model’s sequential forecasting accuracy using only data from weeks 1-4, then weeks 1-8 and then weeks 1-12 finding that a peak in MSM cases in August has been a robust forecast over the first few months of the monkeypox outbreak in the UK (see Fig. S3 and **Supporting information**). We assume that case detection (inferred to be 64% (28%, 96%); table S1) is the same for all MSM groups and non-MSM; lower values in non-MSM where MPX infection is not suspected could lead us to overlook a substantial outbreak among this group. We may also be under-estimating the transmission potential in non-MSM due to the absence of true network structure of the model. However, the likelihood of these oversights diminish as time passes; the sequential forecasts for non-MSM cases early in the outbreak found a substantial outbreak among non-MSM to be plausible, but this projection has diminished the longer the outbreak has stayed relatively fixed in the MSM community (Fig. S3).

The key public health conclusions are: the current case data suggest that monkeypox infection is unlikely to be sustained outside the MSM population (R∼0.07, although this was much higher but still less than early in the outbreak); that public health messaging has already had a significant impact in reducing the risks of transmission by increasing awareness in both MSM and non-MSM populations; finally that rapid vaccine rollout is a high priority to curtail transmission in the most sexually active populations, leading to a more rapid decline in cases.

## Methods

We simulated MPXV transmission in the United Kingdom as a dynamical process where the underlying population at risk was represented as an integer sized, and subdivided by MSM status, sexual activity and a random sized metapopulation. The daily dynamics of the spread of MPXV were encapsulated in a series of stochastic events such as transmission, incubation, and recovery and were assumed to occur stochasticly. Each week a sub-sample of individuals that had their symptom onsets the previous week become reported cases, which connects the underlying transmission model to the observable data.

The design philosophy of this MPXV transmission model was to:

1. Create a sufficiently parsimonious representation of the MPXV transmission structure that Bayesian posteriors for model parameters could be inferred within reasonable time with limited computational resources.
2. Capture the important features of heavy-tailed sexual contact networks, by sub-dividing the MSM population into sexual activity groups by their rate of forming new sexual partnerships.
3. Capture the effect of any additional population structure using a random sized metapopulation subdivision of the MSM population; metapopulation models being known to be reasonable approximations to more detailed individual based models [15].

The modelling approach in this paper is a hybrid of two well-known transmission model types for the spread of MPXV between MSM forming new partnerships. If there was only one metapopulation; then the transmission model used here would be a *random partnership model* [17]. If there was only one sexual activity group then the transmission model used here would be a *metapopulation model* [18]. MPXV spread via other pathways then newly formed MSM sexual partnerships is modelled as a simple homogeneous transmission process.

### Infectious episode progression model

We model the progression of MPXV infection as a **SE**_**n**_**IR** compartmental epidemic model [17]: susceptible individuals (**S**) contract MPXV and are infected without being infectious for the duration of a n-stage process (**E**_**k**_, k = 1…n) before becoming actively infectious (**I**), during which time they can infect other individuals and thus generate further cases. After the actively infectious period individuals recover (**R**) and remain immune to reinfection over the remaining simulation period.

Following the bulk of the MPXV literature, we assume that the latency period (duration between infection and becoming actively infectious) and the incubation period (duration between infection and developing symptomatic disease) are the same; that is that infected individuals are infectious when they show symptoms [4]. A recent study on the incubation period of MPXV estimates a mean incubation period of 8.5-9.6 days [19]; reusing this data and code to make inference on the incubation period as a Gamma distribution of shape and scale parameters κ and θ, respectively, gave mean posterior estimates of 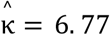 and 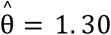. For a daily probability *p*_*inc*_ of progressing between successive stages of our model’s *n*-stage incubation period, the number of days spent in incubation will be given by *n* + *d*, where the distribution *f* of *d* is negative binomial,

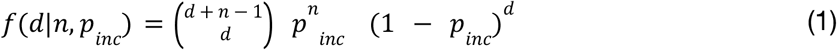

(since *d* is just the number of days in the incubation period where the individual does not progress to the next stage of infection). Using the method of moments to find a negative binomial distribution with identical mean, and minimised difference in standard deviation, to the Gamma distribution inferred from data, we found that a four stage incubation process (i.e. n=4 in the **SE**_**n**_**IR** compartmental structure) with a daily probability of *p*_*inc*_ = 0. 455 of transitioning between successive stages of infection provided an optimal match between our negative binomial model and the Gamma distribution (average incubation period is 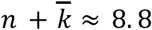 days, with negative binomial mean 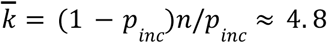, vs Gamma mean 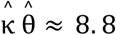 days, negative binomial standard deviation 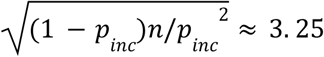 Gamma standard deviation 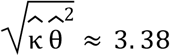, Fig. 3).

**Figure 3.**
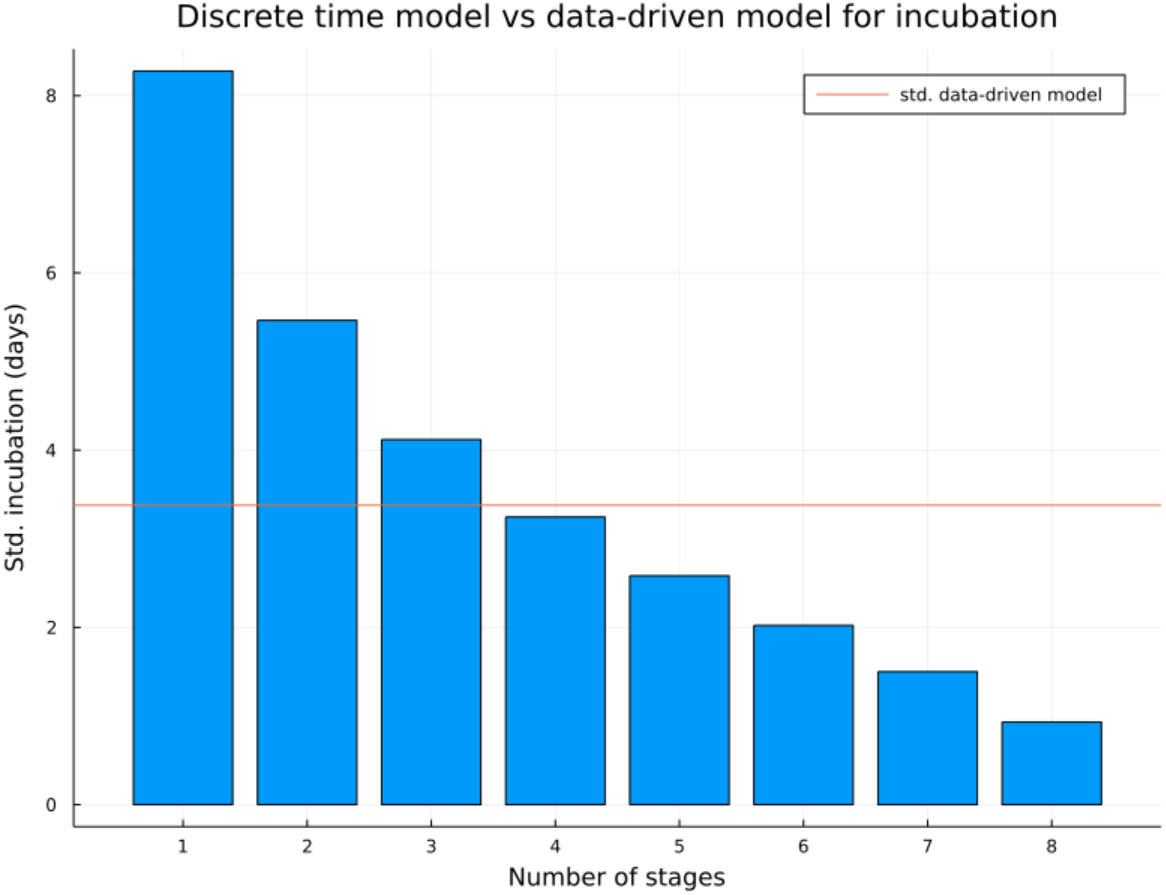
Standard deviation for multi-stage discrete time incubation model when mean is fixed to be 8.8 days by number of stages. The standard deviation for incubation period fitted from data is given as a horizontal red line. The closest match (n=4 stages) was used in simulations.

While it is likely that infected individuals are infectious whilst symptoms persist, which is typically 2-4 weeks [7], we assume that their effective infectious period is potentially self-limiting due to infected individuals reducing their contacts in response to deteriorating MPXV symptoms. We model the infectious period as a one stage process with the mean period μ_*inf*_ being a target for inference; using the same reasoning applied for the total incubation period, the duration of an individual’s infectious period is given by *d* where *d* is negative binomially distributed with parameters 1 and *p*_*rec*_, so that the mean duration is given by μ_*inf*_ = 1 + *p*_*rec*_ /(1 − *p*_*rec*_) = 1/*p*_*rec*_.

### Population structure

We subdivided the population of the United Kingdom (N = 67.2m) into men who are currently actively having new sexual contacts with other men (MSM) and a non-MSM population consisting of the rest of the UK population, including men who identify as MSM but are not currently active or in stable partnership. We further subdivided the active MSM population so that each person belonged to one of 10 sexual activity groups *g* = 1, ..., 10 and one of a random number of metapopulation sub-groups *m* = 1, 2, ....

The proportion of the over 18 year old male population identifying as LGB in the UK has been estimated as 3.4% [20], and the proportion of MSM having at least one new sexual contact with another man in a year, has been estimated as 84.6% [21]. We combine these to make a crude estimate of the size of the sexually active MSM population in the UK, *N*_*msm*_ = 760, 839.

The population distribution of new sexual contacts per year for members of the sexually active MSM community, *k*, has previously been estimated as a power law *f*(*k*) ∼ *k*^−1.82^ [21]. We assumed that the maximum number of new sexual contacts per year was 3650, which defined a proper distribution of yearly sexual contacts on the support of [1,3650]. We divided the MSM population into 10 sexual activity groups by the partitioning the yearly contact rate 1 = *k*_1_ < *k*_2_ <···< *k*_9_ < *k*_10_ = 3650 such that,

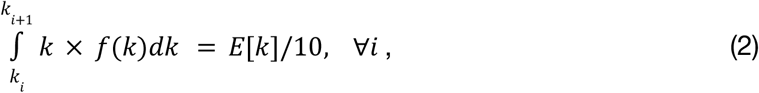

By dividing according to equation (2) the expected rate of new sexual partnerships was equal across groups, which minimises the loss of information in sexual partnership formation implied by discretising the active MSM population. The size of the most active of the 10 sexual activity groups (over the entire metapopulation) was only ∼400 / 760,839 people (∼0.05% of MSM population in UK). In general, this defined both *p*_*g*_ = (*p*_*g*,1_, ···, *p*_*g*,10_), the proportion of MSM typically in each sexual activity group, and, μ_*g*_ = (μ_*g*,1_, ···, μ_*g*,10_), the mean **daily** rate of creating a new sexual contact condtional on being a member of a sexual activity group (Fig. 4).

**Figure 4.**
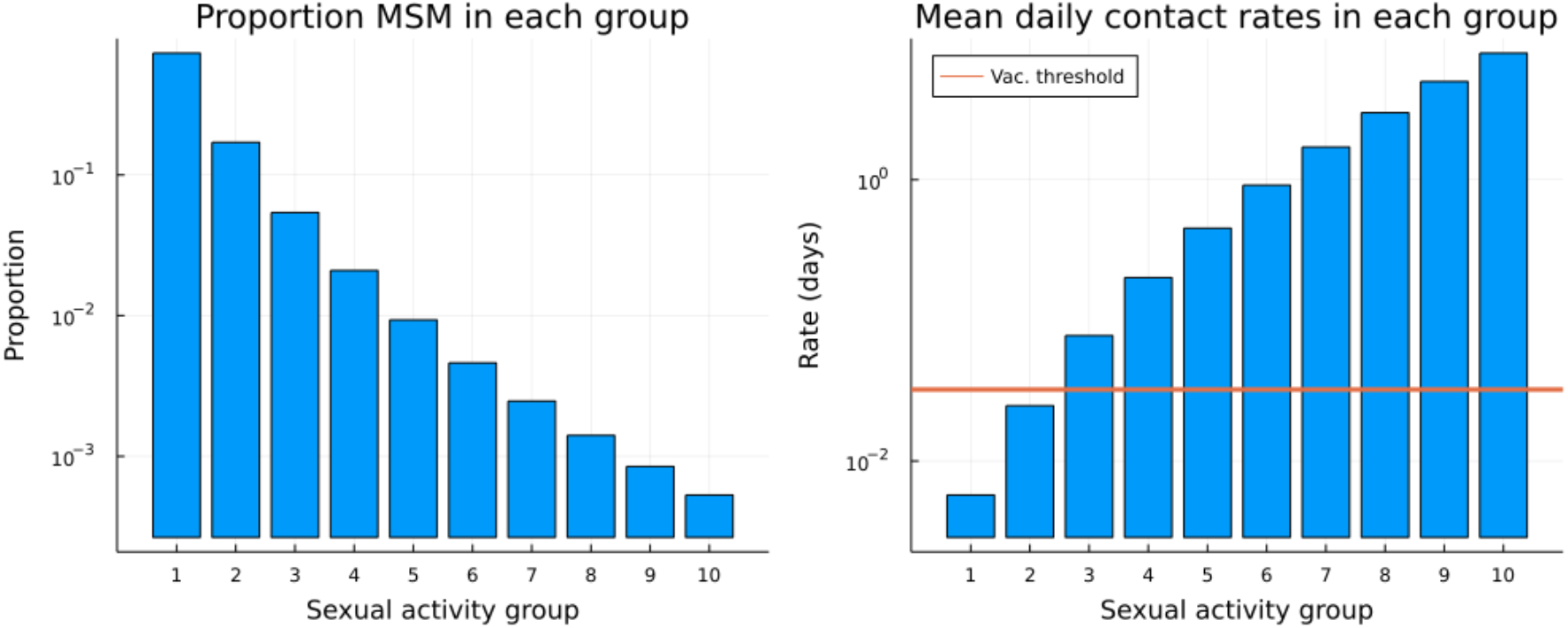
Proportion of MSM in each sexual activity group, and daily rate of new sexual partnership formation. The proportion of the active MSM population in each sexual activity group (*left)*; *NB* this is shown on a log-scale. The mean rate of forming new sexual partnerships by sexual activity group (*right*). Throughout this paper it is assumed that the MSM individuals being offered vaccination are those who typically have at least one new sexual partner per month (*shown as red line*). For this model structure this corresponds to sexual activity groups 3-10 (the 9.4% of the active MSM population most sexually active).

The random sized metapopulation structure was generated according to a Dirichlet multinomial distribution, that is a multinomial where the vector of choice probabilities is drawn from a Dirichlet distribution, *n*_*meta*_ ∼ *Dirichletmultinomial*(*N*_*msm*_, α_*m*_1), where α_*m*_is a real-valued dispersion parameter, 1 is a length 50 vector of ones, and *n*_*meta*_is the resulting vector of metapopulation sizes. Any metapopulation with size 0 was then eliminated from the simulation. It should be noted that α_*m*_→ 0 implies that with probability 1 there will only be one metapopulation of size *N*_*msm*_, whereas in the limit α_*m*_→ ∞ the metapopulation size size distribution is asymptotically multinomial distributed with on average equal sized metapopulations with mean size *N*_*msm*_/50 = 15, 216.

After generating the random sized metapopulations, each metapopulation is further subdivided into sexual activity groups according to a multinomial sample on *p*_*g*_, that is the generated population size of the *g* sexual activity group in the *m* metapopulation (*N*_*g,m*_) is conditionally distributed *N*_*g,m*_|*n*_*meta*_∼*Multinomial*(*n*_*meta*_[*m*], *p*_*g*_).

A new random metapopulation structure was generated for each simulation, with α_*m*_a target parameter for inference (see Parameter Inference).

### Vaccination rollout modelling

UKHSA has secured thousands of vaccines which are being offered to frontline healthcare workers, contacts of cases, and LGB men at highest risk [22]. Within the model we interpret LGB men at highest risk as people within the MSM group who typically have a new sexual contact at least once a month (sexual activity groups 3-10, representing the 9.4% of most sexually active MSM people; Fig. 4).

It has been challenging to capture exact numbers of vaccines that have been taken up by LGB men, however in London (Capital city of the United Kingdom) the reported number of vaccines delivered to MSM people was ∼1000 on the weekend of the 16th/17th July and expected to be 2000 on weekend of 23rd/24th July, and sufficient vaccines had been ordered to offer 5000 doses each weekend in August [23]. In our modelling we assume that the NHS meets these targets, and that an additional 50% of vaccines are accepted by MSM people outside London (Fig. 5). Within the model all vaccines are deployed at the end of each week, and are modelled as becoming effective after a one week delay.

**Figure 5.**
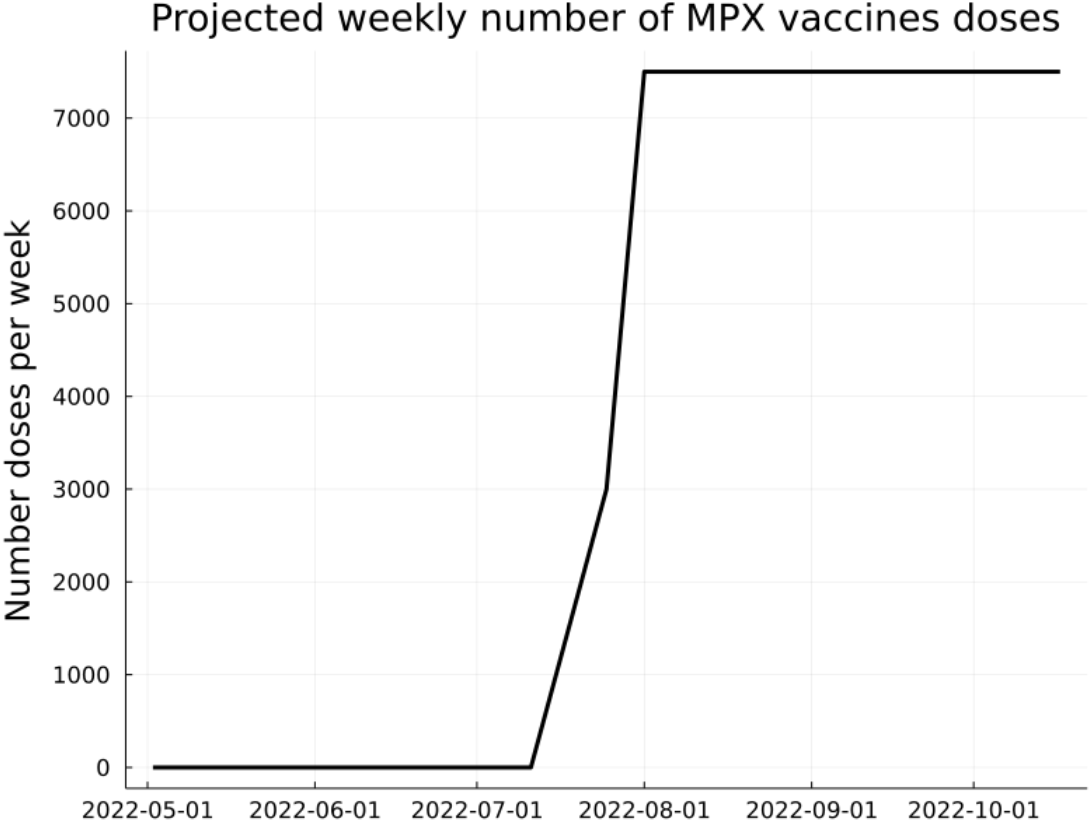
Projected number of weekly vaccine doses given in the United Kingdom.

The effectiveness of smallpox vaccine against monkeypox has been estimated as ∼85% [2,3]. We interpret this as the efficacy of smallpox vaccine against *acquisition* of monkeypox rather than just an endpoint efficacy against disease, although it is not possible to distinguish between the two in the data that is publically available. The proposed dose regime in the UK is to give as many first vaccine doses as possible to LGB men at highest risk, with second doses to be given later as supplies become available [22]. It is possible that vaccination against monkeypox will be less efficacious than 85% against acquisition under this dose regime, therefore whenever modelling the effect of vaccines we draw a vaccine effectiveness parameter *ν*_*eff*_∼ *Uniform*(0. 7, 0. 85) at the start of each simulation.

### Modelling behavioural change due to monkeypox epidemic

Behavioural response and attitude to risk were known to significantly affect the trajectory of the COVID-19 pandemic [13]. We model behavioural change as occurring at two change points:

1. On some date *T*_1_between 1th May 2022 and 18th July 2022 (the change point date is a target for inference);
2. *T*_2_= 23rd July 2022, the date of the announcement by the WHO that monkeypox represented a public health emergency of international concern (PHEIC) [24].

The effect of behavioural change is assumed to reduce transmission for infectious individuals in both the MSM and non-MSM populations through some combination of:

- Voluntary self-isolation during MPX symptoms.
- The effect of any treatments against MPX.
- The effect of contact tracing in raising awareness of their exposure, and guiding safer behaviours.
- The social avoidance of other individuals towards individuals with MPX symptoms.

At the behavioural change points the probability of infection per new sexual contact and the *R*_0_for other routes of transmission decrease by some proportion (see <u>Force of infection</u> below); these proportions of transmission decrease were a target for inference along with the timing of the change point *T*_1_. The United Kingdom Health Security Agency (UKHSA) have not currently found significant evidence of any change in behaviour among identified monkeypox cases in the United Kingdom (personal communication), therefore, 1) our inference priors for the change point at the WHO announcement are towards minimal change (Table S1), and 2) a use a scenario where there is no behavioural response to the WHO announcement (the “no further behavioural change” scenario used in **Results**).

### Force of infection

We consider two transmission pathways for MPX: 1) transmission during close contact when men have sex with men form new sexual partnerships, and 2) all other routes of transmission, including non-MSM sexual partnerships and stable MSM sexual partnerships, household cohabitation and other known transmission pathways for MPXV. The force of infection for each pathway was, respectively,

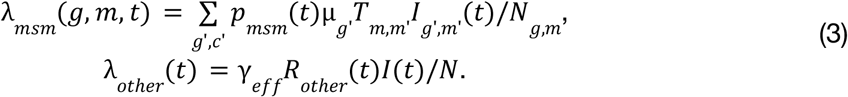

Here *I*_*g,m*_(*t*) is the number of infectious MSM people in sexual activity group *g* and metapopulation *m, I*(*t*) is the the total number of infectious people across the whole population (active MSM and non-MSM), and *N* is the total UK population size. The term *T*_*m,m*’_= 0. 99δ_*m,m*’_+ 0. 01(1 − δ_*m,m*’_) gives cross-metapopulation transmission which defines the implicit meaning of the metapopulation model.

Note that in the limit α_*m*_→ 0, this transmission model recovers ther random partnership model with respect to new sexual formation [17]; this is slightly obscured by the construction of the sexual activity groups, that is that each group has the same rate of sexual contacts overall even though the higher activity groups have far fewer members. Therefore, parameter inference is capable of recreating the random partnership model as the most plausible explanation for spread among active MSM, as well as allowing for more complicated transmission structure within the MSM community if that is a more plausible explanation.

The probability of transmission per sexual contact *p*_*msm*_(*t*) and the reproductive number for other transmission pathways *R*_*other*_(*t*) varied over time because of behavioural change (see <u>Modelling behavioural change due to monkeypox epidemic</u>) as follows:

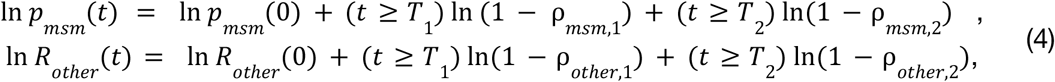

where ρ_*msm*,1_and ρ_*msm*,2_are the proportional risk reductions in MSM sexual contacts at change points 1 and 2 respectively, and ρ_*other*,1_and ρ_*other*,2_are the proportional risk reductions of other transmission pathways at change points 1 and 2 respectively. The daily probability of becoming infected for 1) unvaccinated MSM in sexual activity group *g* and metapopulation *m*, 2) vaccinated MSM in sexual activity group *g* and metapopulation *m* and 3) non-MSM people are:

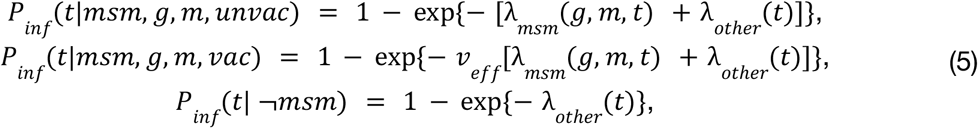

respectively.

### Case detection model

We assume that those MPX cases which are detected have a one week reporting lag after onset of symptoms (i.e. ∼7 days delay with 1-14 day delays possible), which is broadly in-line with the estimated reporting delay in July 2022 [25]. Cases are differentiated by MSM and non-MSM but not by underlying metapopulation or sexual activity group. We modelled the number of cases as being a Beta-Binomial distributed sample over the simulated onsets in the previous week. This is probabilistically equivalent to Binomial sampling but with the probability of detection being independently Beta distributed each week. This is a robust approach to Bayesian inference which reduces the effect of outliers on inference [26], and it has been suggested that stochastic components to detection rate can improve inference in epidemiological modelling by absorbing some of the effect of model misspecification [27], which could be important in this model because in April/May 2022 the reporting delay was probably longer [25]. However, because each week’s detection probability is drawn from an independent Beta distribution our model will not capture temporal directional trends in case detection, for example a trend towards lower chance of detection over time.

The number of MSM and non-MSM cases observed in each week *w* is

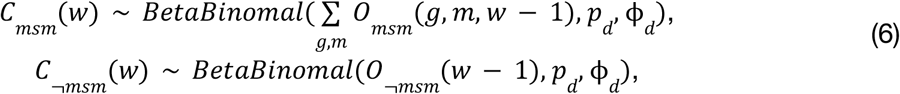

where *O*_*msm*_(*g, c, w* − 1) was the simulated number of symptom onsets in week *w* − 1 in sexual activity group *g* and clique *c* of the MSM population, *O*_¬*msm*_(*w* − 1) was the simulated number of symptom onsets in week *w* − 1 in the non-MSM population, *p*_*d*_is the mean value of the weekly Beta distributed detection rate, and ϕ_*d*_is a dispersion factor for the weekly Beta distributed detection rate. Given *n* onsets in a week the mean and variance in number of cases in the next week are, respectively, *np*_*d*_and *np*_*d*_(1 − *p*_*d*_)(1 + (*n* − 1) ϕ_*d*_). The more common Beta(α, β) parameterization can be recovered via the relationships, *p*_*d*_= α/(α + β) and ϕ_*d*_= 1/(α + β + 1).

### Data and Parameter Inference

Weeks *w* = 1, 2, 3,... were labelled by their Monday date, and all confirmed cases reported were aggregated by week. Confirmed monkeypox case data was retrieved from the *ourworldindata* repository [28], and based on UKHSA reporting [16] we used a stable 95.6% of cases in the MSM population. While this implied that the weekly case data for MSM and non-MSM, *C*_*msm*_(*w*) and *C*_¬*msm*_(*w*) respectively for *w* = 1, 2, 3... were not necessarily integer valued, the error measure we used in our inference [see Equation (7)] did not require integer reference data.

We performed Bayesian inference on the model parameters (see Table S1 for full list of parameters, priors used and posterior mean and 95% CIs) using sequential Monte Carlo based approximate Bayesian computation (SMC-ABC [29]) implemented in the Julia language package ApproxBayes.jl [30]. Forward simulations were performed by solving the stochastic monkeypox transmission model using the DifferentialEquations.jl ecosystem of dynamical system solvers for the Julia programming language [31].

After drawing model parameters from the prior distributions, simulations were initialised at the beginning of the week immediately previous to the week with first reported cases (Monday 25th April 2022, *w* = 0) as follows:

1. A random metapopulation and sexual activity group distribution for MSM people was generated (see Population structure).
2. One metapopulation was randomly selected proportionally to metapopulation size.
3. For the selected metapopulation a Poisson distributed number of individuals were assigned to each incubation stage and the infectious stage (uniformly likely) in each sexual activity group (proportional to population frequency) such that conditional on the chosen *p*_*d*_and μ_*inf*_parameters the expected number of MSM cases on week *w* = 1 was ι_0_, which was a target parameter for inference.

During each simulation a predicted (integer) number of reported cases for MSM and non-MSM on each week was generated using equation (5): *Ĉ*_*msm*_(*w*), *Ĉ*_¬*msm*_(*w*) for *w* = 1, 2, 3, The error metric for the simulation used by the SMC-ABC algorithm against the true data was, *d*_1_(*Ĉ, C*) defined as:

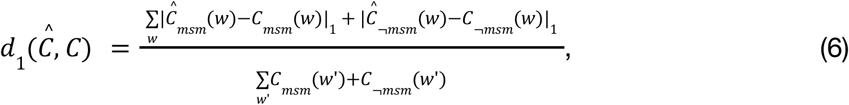

where |.|_1_denotes L1 norm. The last week of available case data was not used in inference because of potentially confounding right-censoring.

Before running SMC-ABC we performed prior predictive model checking and simulation-based calibration for the error target value (see Supporting Information).

## Data Availability

All data for reproducing this work is available online at https://github.com/SamuelBrand1/MonkeypoxUK

## Code availability and ongoing projections

All code and data used in running the model is available at the open github repository: https://github.com/SamuelBrand1/monkeypoxUK. As more data becomes available we will be periodically updating this repository with the most recent projections.

## Acknowledgements

As mentioned in the main document, this work has benefitted from conversations with the UK Health Security Agency (UKHSA) MPX modelling cell and technical committee.

