## Supplementary text and figs for "The role of vaccination and public awareness in medium-term forecasts of monkeypox incidence in the United Kingdom"

<u>Prior predictive checking and simulation based calibration</u>

Following a Bayesian workflow [32] we first generated 1000 parameter samples from the prior distribution $\pi$: $\theta^{(i)} \sim \pi$ for $i = 1,..., 1000$. Then for each parameter sample we simulated a trajectory for weekly MSM and non-MSM cases $\hat{C}^{(i)}_{msm}(w)$, $\hat{C}^{(i)}_{\neg msm}(w)$, $i = 1,..., 1000$. These simulations from the prior distribution were visually inspected to assess the coherence of the prior beliefs, as represented by priors on parameters, with our prior belief on the trajectory of MPXV in the United Kingdom. This demonstrated a reasonably wide spread of *a priori* potential outcomes, including early disappearance, and the eventual domination of the epidemic by non-MSM cases (Fig. S1).

Before running the SMC-ABC inference we generated a target error threshold for accepting a parameter into an empirical ensemble approximating the Bayesian posterior distribution using simulation based calibration. The procedure was to resample a trajectory for weekly MSM and non-MSM cases for each of the prior draws $\theta^{(i)}$: $\tilde{C}^{(i)}_{msm}(w)$, $\tilde{C}^{(i)}_{\neg msm}(w)$ and generate an ensemble of error measures: $d_1(\hat{C}^{(i)}, \tilde{C}^{(i)})$ for $i = 1,..., 1000$. This ensemble represented a spread of typical errors between case trajectories even when both trajectories were generated using the same underlying parameters. The target error value for ABC-SMC parameter acceptance, $\epsilon_{target}$, was chosen to be at the 5th percentile of the generated error measure ensemble (Fig. S1).

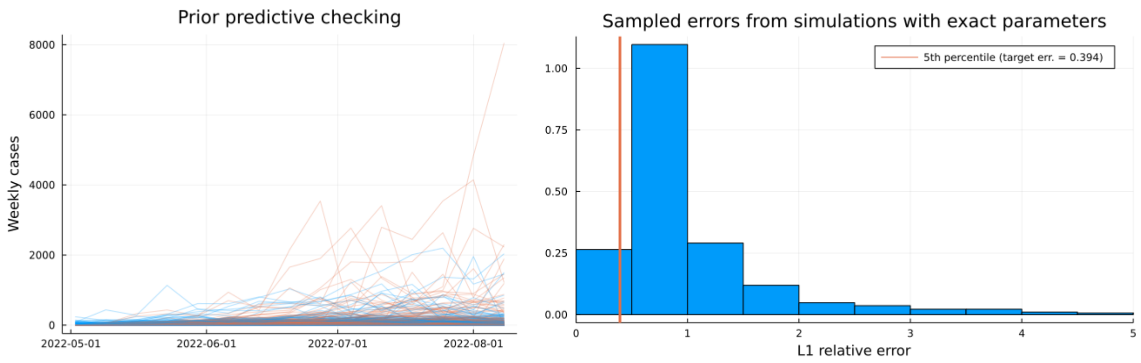


**Figure S1. Prior predictive checking and simulation based calibration.** Trajectories of weekly cases for MSM (*blue lines*) and non-MSM (*red lines*) as generated using the prior distribution on parameters (*left*). The distribution of errors when resampling trajectories using matched parameters with the 5th percentile (*red line*) chosen as the SMC-ABC inference threshold (*right*).

Posterior distribution

| Parameter | Posterior mean (95% CI) | Prior distribution | Description |
|---|---|---|---|
| Population structure and initial condition parameters | | | |
| $\alpha_m$ | 1.06 (0.038, 3.34) | Exp(1) | Dispersion parameter for metapopulation sizes. |
| $\iota_0$ | 1.30 (0.19, 3.72) | LogNormal$(0, 1)$ | Scale parameter for number of initial infected people. |
| Detection probability parameters | | | |
| $p_d$ | 0.65 (0.30, 0.95) | Beta(5,5) | Mean weekly probability of case detection. Weekly probability of detection was random with $P_w \sim Beta(\alpha, \beta)$ with $E[P_w] = p_d$. |
| $\phi_d = 1/(M + 1)$ | 0.09 (0.028, 0.31) | $M \sim Gamma(3, 10/3)$ | Dispersion of weekly probability of case detection ($M$ is the effective sample size $\alpha + \beta$ for the weekly $P_w \sim Beta(\alpha, \beta)$ distributed probability of case detection). |
| Baseline transmission parameters | | | |
| $\mu_{inf}$ | 13.06 days (5.49,21.17) | $(\mu_{inf} - 1) \sim$ Gamma(3,2), conditioned on being in [0,21] days | Mean number of infectious days. |
| $p_{msm}(0)$ | 0.31 (0.15, 0.52) | Beta(1,9) | Baseline probability of transmission per sexual contact |
| $R_{other}(0)$ | 0.47 (0.052, 1.08) | LogNormal($\ln 0.75$,0.25) | Baseline reproductive number non-MSM sexual contacts |
| Behaviour and risk change point parameters | | | |
| $T_1$ | 2nd June (16th May, 30th June) | Uniform(15th May, 18th July) | Change point time for reduction in transmission due to awareness of MPX. |
| $\rho_{msm,1}$ | 30.39% (2.81%, 64.69%) | Beta(1.5,1.5) | Reduction in probability of transmission per |

| Parameter | Posterior mean (95% CI) | Prior distribution | Description |
|---|---|---|---|
| | | | sexual contact after change point at $T_1$. |
| $\rho_{other,1}$ | 81.78% (48.21%, 99.17%) | Beta(1.5,1.5) | Reduction in non-MSM sexual contact reproductive number after change point at $T_1$. |
| $\rho_{msm,2} = s_{msm,2}\rho_{msm,1}$ | 8.92% (0.34%, 26.89%) | $s_{msm,2}$ ~Beta(1,4) | Reduction in probability of transmission per sexual contact after WHO announcement of PHEIC. Parameterized as relative to the effect at the change point at $T_1$ |
| $\rho_{other,2} = s_{other,2}\rho_{other,1}$ | 13.46% (0.34%, 50.93%) | $s_{other,2}$ ~Beta(1,4) | Reduction in non-MSM sexual contact reproductive number after WHO announcement of PHEIC. Parameterized as relative to the effect at the change point at $T_1$ |

**Table S1: Definition of the target parameters for inference and their posterior estimates.**

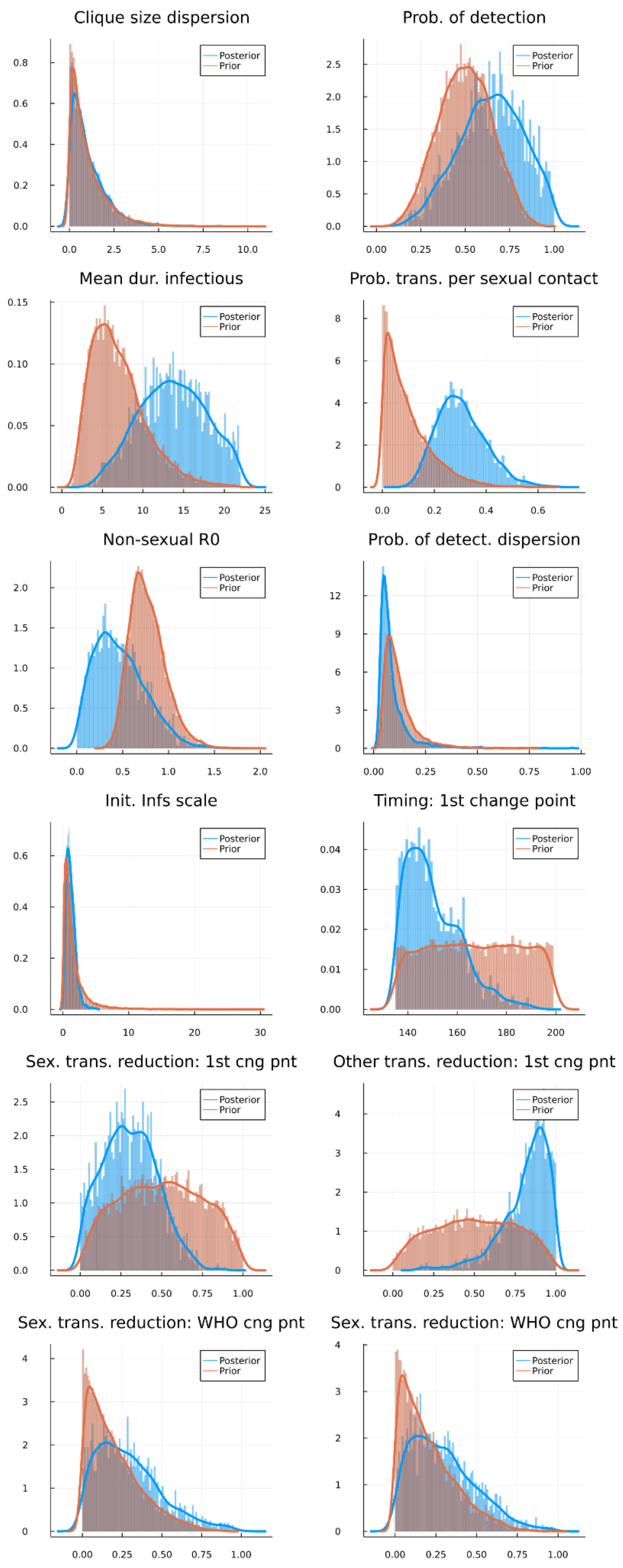


**Figure S2. Empirical distributions of the target parameters for inference (blue and orange colours, posterior and prior distributions, respectively).**

Sequential predictions using redacted data

To assess the predictive quality of the model we refitted using case data only available in the first 4, 8, and 12 weeks, and generated predictions using the Forecast scenario (Fig. S3). In particular, week 8 was an outlier in the case trend over 500 determined MPXV cases, and we were interested to determine the robustness of the model prediction to outlier weeks. We found that in each sequential projection the posterior mean for cases among MSM people peaked in August, which seems to be a robust conclusion of the model. For non-MSM cases, fitted only on weeks 1-4 and weeks 1-8 data, the model posterior predictions are highly uncertain and there are sufficient trajectories with a sustained outbreak among non-MSM people that the posterior mean prediction is 100s of non-MSM cases per week (Fig. S3). By week 12 the inference has shifted the posterior likelihood of a significant outbreak among non-MSM people to negligible and forecasts less than 50 non-MSM cases a week going forwards.

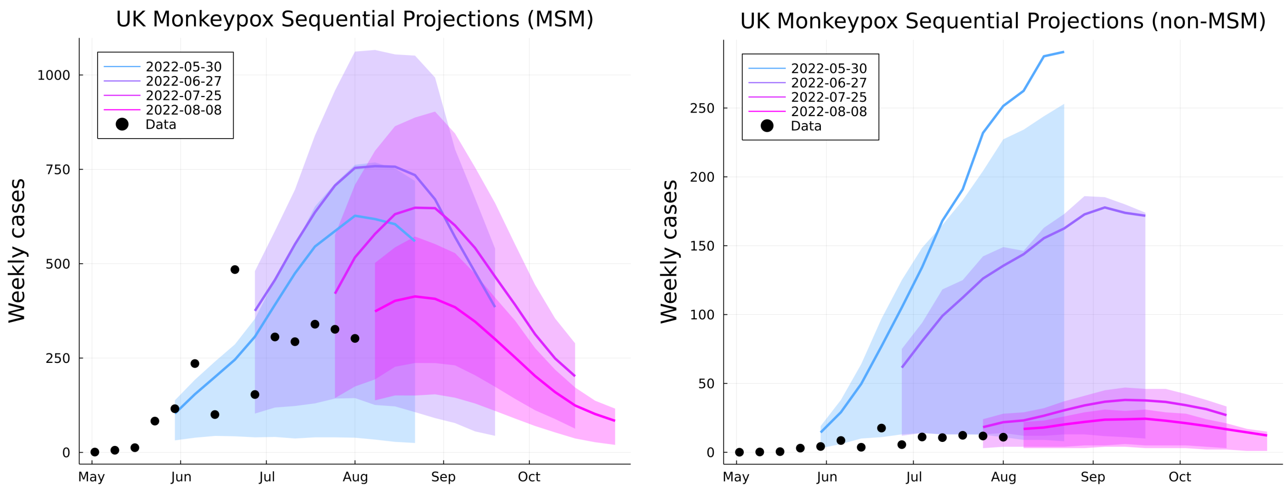


**Figure S3. Sequential predictions of weekly reported cases.** On a scale of blue - purple for increasing weeks of data, the forecast scenario projections for MSM (*left*) and non-MSM cases (*right*). Solid lines are posterior mean predictions, background shading is posterior IQR. NB: because the outbreak sizes were highly skewed, the posterior mean for non-MSM cases trends higher than the 75% posterior probability for weekly case numbers when only week 1-4 data was used for inference.
